# Molecular characterization of antimicrobial resistance organisms from drinking water and wastewater in a metropolitan city

**DOI:** 10.1101/2024.04.12.24305711

**Authors:** Khursheda Akhtar, Nasreen Farhana, Alamgir Hossain, Fahmida Khanam

## Abstract

**Background:** Antimicrobial resistant (AMR) organisms in environment may harm people. This study assessed the phenotypic and genotypic characteristics of AMR organisms from drinking and wastewater.

**Materials and methods:** This cross-sectional study conducted randomly on 30 samples (15 drinking water samples from household places; 15 sewage lifts stations) and collected aseptically, filtered, inoculated and isolated from culture plates, identified biochemically of pathogenic bacteria, and disc diffusion tested for antibiotic susceptibility. The primers of the targeted antimicrobial resistance genes were used for molecular amplification.

**Results:** Twenty-five bacteria were isolated from 30 drinking and wastewater samples. *Pseudomonas* spp. (36%), *Enterobacter* spp. (28%), *Escherichia coli* (20%), *Citrobacter* spp.(4%)*, Acinetobacter* spp (4%) and *Klebsiella oxytoca* (4%) were isolated. Most of the isolates exhibited resistance to multiple groups of antibiotics, with meropenem, imipenem, gentamicin, ciprofloxacin, and amikacin showing the highest sensitivity against the isolates. Multiplex PCR confirmed the presence of two ESBL genes (*bla*SHV and *bla*CTX-M-15) and five carbapenemase genes (*bla*IMP, *bla*VIM, *bla*KPC, *bla*OXA1, *bla*NDM1) in resistant bacteria and *bla*CTX-M-15 (53.3%) followed by *bla*KPC (46.7%) genes were the most prevalent from drinking and wastewater samples respectively. According to WHO’s sanitary inspection risk score classification, 60% of drinking water samples scored 4 out of 11, categorizing them as intermediate risk based on hazard score.

**Conclusions:** This study addresses antimicrobial resistance in the environment, emphasizing public health implications; advocating for improved environmental regulations to mitigate AMR organism discharge through wastewater and drinking water.

**The significance of the study:** The study attempted to determine the pattern of antimicrobial resistance of microorganisms using phenotypic and genotypic methods by polymerase chain reaction (PCR), targeting particular genes with specific sequence of primers. As in Bangladesh very few studies for antimicrobial resistance organisms from drinking water and wastewater around household and hospital environments in Dhaka city, yet finished to change public health perspectives, and inform respective authorities for making decision. Thus the research contributes to generating some evidence-based information about the reservoir of antimicrobial resistance in environment.

## Introduction

Antimicrobial resistance (AMR) has emerged as one of the most serious public health issues of the twenty-first century, threatening the effective prevention and treatment of an ever-increasing range of infections caused by bacteria, parasites, viruses, and fungi that are no longer susceptible to common antimicrobials including antibiotics, antivirals, antifungals and anti-parasites.^1^ The World Health Organization (WHO) has long recognised the need for an improved and coordinated global effort to contain AMR. In 2001, the WHO Global Strategy for Containment of Antimicrobial Resistance has provided a framework of interventions to slow the emergence and reduce the spread of antimicrobial-resistant microorganisms.^2^ In 2012, WHO published The Evolving Threat of Antimicrobial Resistance – Options for Action proposing a combination of interventions that include strengthening health systems and surveillance; improving use of antimicrobials in hospitals and in community; infection prevention and control; encouraging the development of appropriate new drugs and vaccines; and political commitment.^3^

During the last years, the importance of the environment in the spread of antibiotic resistance has been widely recognised. Water is a major way of dissemination of bacteria between different environmental compartments. Infectious diseases caused by pathogenic bacteria, viruses and parasites (e.g. protozoa and helminths) are the most common and widespread health risk associated with drinking-water.^4^ Large amounts of antibiotics are released into municipal wastewater due to incomplete metabolism in human beings or due to disposal of unused antibiotics. Some available data show that antibiotic resistant bacteria and antibiotic-resistant genes can be detected in wastewater samples and that the conditions in wastewater treatment plants (WWTPs) are favourable for the proliferation of resistant bacteria.^5^

The public health burden is determined by the severity and incidence of the illnesses associated with these pathogens, their infectivity and the population exposed. The threat AMR poses on global public health necessitates mitigation of dissemination of AMR bacteria, and therefore the identification of possible dissemination routes.^6^ Breakdown in water supply safety (source, treatment and distribution) may lead to large-scale contamination and potentially to detectable disease outbreaks. In some cases, low-level, potentially repeated contamination may lead to significant sporadic disease, but public health surveillance is unlikely to identify contaminated drinking-water as the source.^4^

AMR occurs when bacteria, viruses, fungi and parasites change over time and no longer respond to medicines making infections harder to treat and increasing the risk of disease spread, severe illness and death. The main drivers of antimicrobial resistance include the misuse and overuse of antimicrobials; lack of access to clean water, sanitation and hygiene (WASH) for both humans and animals; poor infection and disease prevention and control in health-care facilities and farms; poor access to quality, affordable medicines, vaccines and diagnostics; lack of awareness and knowledge; and lack of enforcement of legislation.^7,8^

The One Health approach brings together multiple sectors and stakeholders engaged in human, terrestrial and aquatic animal and plant health, food and feed production and the environment to communicate and work together in the design and implementation of programmes, policies, legislation and research to attain better public health outcomes.^9^

Currently, resistant infections result in 700,000 deaths every year, but the global resistance-associated mortality is estimated to top 10 million lives per year in 2050. Along with high income countries (Europe, USA); in low- and middle-income countries, AMR infections have been responsible for the deaths of 58,000 children and 38,000 adults in India and Thailand, respectively.^10^

Antimicrobial resistance (AMR) has become an emerging issue in the developing countries as well as in Bangladesh. A study was conducted by Islam MA (2017) that wastewater samples from hospital-adjacent areas (HAR) and from community areas (COM), as well as public tap water samples, for the occurrence and characteristics of New Delhi metallo -1(NDM-1) producing bacteria. Of 72 HAR samples tested, 51 (71%) samples were positive for NDM-1-producing bacteria, as evidenced by phenotypic tests and the presence of the *bla*NDM-1 gene, compared to 5 of 41 (12.1%) samples from COM samples (P 0.001). *Klebsiella pneumoniae* (44%) was the predominant bacterial species among *bla*NDM-1-positive isolates, followed by *Escherichia coli* (29%), *Acinetobacter* spp. (15%), and *Enterobacter* spp. (9%).^11^

These bacteria were also positive for one or more other antibiotic resistance genes, including *bla*CTX-M-1 (80%), *bla*CTX-M-15 (63%), *bla*TEM (76%), *bla*SHV (33%), *bla*CMY-2 (16%), *bla*OXA-48-like (2%), *bla*OXA-1 (53%), and *bla*OXA-47-like (60%) genes.^12^ In another study by Toleman (2012) found that, in Bangladesh, the *bla*NDM-1 gene were in 62% of environmental waters and in fermentative and non-fermentative gram-negative bacteria. Resistance genes and plasmid profiles for a subset of Escherichia coli strains was found in a study of extensively drug-resistant NDM-1–encoding bacteria in the environment, Dhaka, Bangladesh.^13^ The presence of NDM-1 β-lactamase-producing bacteria in environmental samples in New Delhi has important implications for people living in the city who are reliant on public water and sanitation facilities.^14^

As previously discussed, AMR is driven by complex interacting factors that could be described as a resistance network.^15^ This network forms links between clinical factors (e.g., human health, animal husbandry and veterinary medicine) and other components, including human activities (e.g., travel, human displacement and over and misuse of antimicrobial drugs) and environmental factors (e.g., persistence of antimicrobial drugs and AMR organisms in soil and water).^16–20^

The selective pressure associated with the exposure to antimicrobials in healthcare, agriculture and the environment enhances the development of new AMR variants and novel resistance mechanisms.^21^ Transmission of AMR can spread between people, animals and the environment via a number of different routes.^22^ The environment acts as a bridge for different compartments, between animals to compost to soil to water to sediments to sewage.^23^ While the environment acts as the reservoir, it also works simultaneously to mix mobile genetic elements (MGEs) that interact and diffuse into other parts or into human and animal hosts.^22, 24,25^

Non-judicial use of antibiotics is considered to be most significant reason for the emergence selection and spreading of antibiotic-resistant bacteria in both animals and humans. Via food and water antibiotics can lead to the emergence and dissemination of different resistant bacteria. Wastewater contains the faecal material with E coli; simultaneously wastewaters are the prominent source of antibiotic resistance bacteria. In Dhaka city 30 % sewage are only treated rests 70% are untreated and excrete into environment, and hospital waste as well. There is limited information about the Extended-Spectrum Beta Lactamase (ESBL) gene carriage in Bangladesh in environment particularly in drinking and wastewater.^26^ AMR is a complex problem that requires a united multi sectorial approach. Without AMR containment, the Sustainable Development Goals for 2030-such as ending poverty, ending hunger ensuring healthy lives, reducing inequality and revitalizing global development partnerships are less likely to be achieved.^27^

It is critical to strengthen and harmonize the AMR surveillance through the development of agreed epidemiological and microbiological methods, the adoption of common definitions to enhance the ability to share and compare resistance information, and to attain a better coordination of the surveillance networks. To identify the magnitude of problem, this is also true for policies to prevent contamination of the environment with antimicrobial residues from human and animal use.

## Objectives

### General objective

To detect the pattern of organisms in wastewater and drinking water with the extent of their antibiotic resistance and presence of resistance genes

### Specific objectives

1. To isolate and identify of pathogenic organisms in drinking and wastewater
2. To detect antimicrobial resistant organisms phenotypically [by double disc synergy test (DDS), combined disc (CD) assay, and modified Hodge test (MHT)] from drinking and wastewater
3. To identify antimicrobial resistance gene by multiplex PCR (genotype encoding *bla*SHV, *bla*CTX-M15 genes for beta-lactamases producers and *bla*OXA1, *bla*NDM-1, *bla*VIM, *bla*IMP and *bla*KPC genes for carbapenemase producers)

## Rationale

Antimicrobial resistance (AMR) is the major issue posing a serious global health threat. Low- and middle-income countries are likely to be the most affected both in terms of impact on public health and economic burden.^27^ The cost of AMR to the economy is significant. In addition to death and disability, prolonged illness results in longer hospital stays, the need for more expensive medicines and financial challenges for those impacted. Without effective antimicrobials, the success of modern medicine in treating infections, including during major surgery and cancer chemotherapy, would be at increased risk. Recently, the World Health Organization (WHO) Global Antimicrobial Surveillance System (GLASS) reported increased levels of resistance in a number of serious and common bacterial infections in many regions of the world.^29^

Anthropogenic activities, hospital effluents, industrial wastes, domestic sewage, and urban/agricultural runoffs represent a significant source of emerging biological contaminants including pathogenic organisms, antibiotic residues, antibiotic resistant bacteria (ARB) and antibiotic resistant genes (ARGs) in aquatic ecosystems.^30^ The uncontrolled use of antibiotics exposes bacteria to antibiotics, which leads to subsequent acquirement of resistance through mutations or by eliminating nonresistant bacteria.^31^ The overuse of antibiotics and their subsequent poorly managed release to the environment have been linked with the development of such undesired characteristics. Hospitals and animal farms are amongst the leading contributors of antibiotics, bacteria and antibiotic resistance genes and toxic metals in the environment.^32^ However, the situation is a lot worse in developing countries lacking adequate infrastructures to contain the spread of contaminated waters and where wastewaters are rarely treated.

The selective pressure associated with the exposure to antimicrobials in healthcare, agriculture and the environment enhances the development of new AMR variants and novel resistance mechanisms. Transmission of AMR can spread between people, animals and the environment via a number of different routes. While the environment acts as the reservoir, it also works simultaneously to mix mobile genetic elements (MGEs) that interact and diffuse into other parts or into human and animal hosts.^33^ Beta-lactams are most widely used antibiotics including natural and synthetic penicillin and their derivatives; resistance to beta-lactam antibiotics is of special concern for human being.^34^ Human infection due to ESBL producing bacteria is associated with increased mortality, morbidity, high cost of hospitalization and delay appropriate therapy. *Escherichia coli* and *Salmonella spp.* are often common ESBL producers isolated in environment. These bacteria are resistant to penicillins, cephalosporins and aztreonam mainly due to the production of CTX-M, TEM and SHV β-lactamases, which are encoded by *bla*CTX-M, *bla*SHV, and TEM genes respectively.^35^ There is limited information about environmental factor especially waste and drinking water on ESBL/MRSA gene carriage in Bangladeshi.

Thus the research contributes to generating some evidence-based information about the reservoir of antimicrobial resistance in environment. The implications of our findings are likely to be significant from a public health standpoint. The urgent need to place efficient wastewater treatment plants in health care settings as part of biosecurity programs. More work should be done to understand the spread of MDR bacteria, including the ESBL/NDM-1-producing ones in the environment and the resultant health implications. In future with these samples further research (genomic sequencing) was done.

## Methodology

This was a cross sectional study on environmental samples (drinking water from household and wastewater from sewage lift stations near hospitals) in Dhaka city at National Institute of Preventive and Social Medicine, Mohakhali, Dhaka, Bangladesh. A total 30 samples was collected, among them 15 samples of wastewater and 15 samples of drinking water. In Dhaka city, WASA distribute the service by 10 MODS zone^36^ and from these zones, 5 MODS zones were selected randomly. For wastewater, 10 sample of wastewater randomly select from 28 sewage lift stations in Dhaka city and five (05) samples purposively selected from sewage lift station near the hospitals (wastewater drains within 3 mile of boundaries of the large public/private hospitals or clinics). For drinking water, 15 samples were selected from 15 point of use (household reservoirs/ public water supply that local residents use for drinking, washing, and food preparation) randomly by using Google map. A structured questionnaire and checklist was approached for sample collection and laboratory purpose.

### Sample collection procedure

The sample from a drinking tap or wastewater were collected after maintaining aseptic precaution according to standard guidelines.^37^ Immediately after collection, samples were placed in an insulated ice boxes for transport to NIPSOM laboratory, Mohakhali, Dhaka. Water samples were examined as soon as possible on arrival and always within 6 hours of collection. Long distance sample was dipped into Cary *Bla*ir transport medium (Hi Media Laboratories, Mumbai, India) before shifting into the laboratory.

### Bacterial isolation and identification

The water samples were filtered with 0.02 micron filter paper and then inoculated in 5mL of nutrient broth and incubated overnight at 37°C and the swabs from the suspected growth of the broths were then inoculated in non-selective culture plates and incubated again overnight at 37◦C.^38^ The isolated colonies was further streaked on various differentiation agar media (i.e. MacConkey agar, Chromogenic Coliform Agar) for selective bacterial isolates. All the isolated organisms were identified by their colony morphology, hemolytic criteria, staining characters. Gram stained smear was examined for presence and arrangement of the organisms for presumptive identification and to correlate these findings with results of biochemical tests. Gram positive isolates was also differentiated by arrangement in Gram staining, colonial characteristics, catalase test and coagulase tests. Gram negative bacteria was identified based on colonial morphology and pigmentation, oxidase test, growth at 42°c, carbohydrate fermentation, H_2_S production, citrate utilization, motility, indole formation, and urea hydrolysis.^37^

### Antimicrobial sensitivity test

This test was performed by Kirby-Bauer modified disc diffusion method on Mueller-Hinton agar media according to the standards published by the Clinical and Laboratory Standards Institute (CLSI) and inoculated plates was incubated aerobically at 37°C for 24 hours. After 24 hours incubated each plate was examined to ensure the confluence growth of organism^39^ and evaluated with standard chart of CLSI guideline into sensitive, intermediate and resistant categories on the basis of zone of inhibition around the disc.^40^ To assess the performance of the method, *Esch. coli* ATCC 25922 was used as a control strain. ^39^

### Phenotypic detection of antimicrobial-resistant bacteria

Antimicrobial-resistant Gram positive bacteria was phenotypically detected with standard disc diffusion method as per CLSI standards i.e. Methicillin resistance for *staphylococcus aureus* (MRSA) using oxacillin (1µg) and cefoxitin (30 µg) disc, vancomycin resistant enterococci (VRE) using vancomycin (30 µg) disc and extended spectrum beta lactamases (ESBL) producing Gram negative bacilli and carbapenemase producing enterobacteriaecae (CRE) by double disc synergy test (DDS), combined disc (CD) assay, and modified Hodge test (MHT) respectively.^41,42^

### Molecular characterization of antibiotic-resistant bacteria

The conventional polymerase chain reaction (PCR) method was applied for molecular characterization and freshly cultured isolates bacteria was used to prepare template deoxyribonucleic acid (DNA) by the boiling method.^43^ The bacterial samples were grown on nutrient agar plates and a single colony was inoculated in the nutrient broth. The cultures was grown overnight and centrifuged at 5000 rpm for 5 min at room temperature and the bacterial pellet was washed with Tris buffer (1M Tris-hydrochloride [HCl], 0.1M ethylenediaminetetraacetic acid [EDTA] and 0.1 M sodium chloride [NaCl]). The pellet was suspended in 300 µl of sterile distilled water into eppendorf tubes and vortexed until mixed. The tubes were heated at 100⁰C for 10 minutes in a heat block (DAIHA Scientific, Seoul, Korea). After heating, immediately the eppendorf tubes were placed on ice for 5 minutes and then centrifuged at 14,000 g at 4⁰C for 6 minutes. The supernatant were taken into another micro centrifuge tube by micropipette and were used for DNA template. The extracted DNA was preserved at 4⁰C for 7-10 days and -20⁰C for long time.^44^ The sequences of designed primers encoding *bla*SHV, *bla*CTX-M-15 and *bla*OXA-1 genes for beta-lactamases producers and *bla*NDM-1, *bla*VIM, *bla*IMP and *bla*KPC genes for carbapenemase producers^27,28^. downloaded from the GenBank database was used in this study (Table 2). Primers were mixed with Tris-EDTA (TE) buffer according to manufacturer’s instruction. PCR was performed in a final reaction volume of 25µl in a PCR tube, containing 10 µl of mastermix [premixed mixture of 50 units/ml of Taq DNA polymerase supplied in a proprietary reaction buffer (pH 8.5), 400μM dNTP, 3mM MgCl_2_ (Promega corporation, USA), 1 µl forward primer and 1 µl reverse primer (Promega corporation, USA), 3 µl extracted DNA and 14 µl nuclease free water for monoplex PCR. After a brief vortex, the PCR tubes were centrifuged in a micro centrifuge machine for few seconds. PCR assays were performed in a DNA thermal cycler. Each cycle of PCR reaction consists of initial denaturation at 95°C for 10 minutes followed by annealing temperature which varied in different reaction followed by elongation at 72°C for 1 min. After 35 cycles final extension was done at 72°C for 10 minutes. Then the product was held at 4°C. After amplification, products were processed for gel documentation by electrophoresis on 1.5% agarose gel. After electrophoresis, the gel was stained with ethidium bromide (10 µl ethidium bromide in 100 ml distilled water) for 30 minutes. It was then de-stained with sterile distilled water for 15 minutes. The gel was then removed from the tray and observed under UV Transilluminator (Gel Doc, Major science, Taiwan) for DNA bands. The DNA bands were identified according to their molecular size by comparing with the molecular weight marker (100 bp DNA ladder) loaded in a separate lane. Samples showing the presence of corresponding bp band were considered positive for the presence of that organism. Exact band size was calculated from a log graph.^42–48^

### Statistical analysis

Data was verified, entered and subsequently analyzed using Statistical Package for Social Science (SPSS) software of recent version. Then, responses were initiated with descriptive analysis. Frequencies, percentage, mean, median, mode, range, and standard deviation were calculated for continuous variables, while frequencies and percentages were calculated for categorial variables. Tables and graphs were used to present the data.

### Ethical considerations

Prior to doing data collection, ethical clearance was obtained from Ethical review committee of BMRC (Memo no.: BMRC/HPNSP-Research Grant/2022-2023/613(1-18), Date: 15.01.2023). Permission was taken from the respective authority (DWASA)(Memo no.- 46.113.519/520.00.00.154.2023.443.mi.chem.dept.; Date:29.03.2026) for wastewater collection and a signed informed consent was taken from the household convincing that privacy was maintained. For safeguarding confidentiality and protecting anonymity each of the samples was given a special ID no. which was followed in collection, transport to lab and reporting, in each and every step of the procedure.

## Results

### Isolation pattern of pathogenic bacteria

A total of 19 (63.3%) growths were yielded from the culture of 30 water samples examined. Among them, four (26.7%) isolates out of 15 drinking water and 15 (100%) isolates from all the wastewater samples were detected (Table 1). The drinking water yielded single organism from 3 (20%) of samples while one (6.7%) sample was identified with more than two organisms. On the other hand, the wastewater yielded single organism from twelve (80.0%) samples, two organisms from two (13.3%) samples and more than two organisms were detected from one (6.7%) samples (Table 2). A total of 25 organisms were isolated from 30 samples of both drinking and wastewater and the predominant bacteria were *Pseudomonas* spp. (36%) followed by *Enterobacter* spp. (28%), *Escherichia coli* (20.0%), *Citrobacter freundii* (4.0%)*, Citrobacter koseri* (4.0%)*, Acinetobacter* spp (4.0%) and *Klebsiella oxytoca* (4.0%) (Table 3). The isolated pathogenic bacteria from drinking and wastewater samples were then identified by relevant biochemical tests (Table 4 and 5)

**Table 1:** Growth on culture from water samples (N=30)

| Water samples | No growth<br>f (%) | Growth<br>f (%) |
| --- | --- | --- |
| Drinking Water<br>(n =15) | 11 (73.3) | 4 (26.7) |
| Wastewater<br>(n =15) | 0 | 15 (100) |
| Total (N=30) | 11 (36.7) | 19 (63.3) |

**Table 2:** Pattern of bacterial growth isolated from water samples.

| Water samples | Single organism<br>f (%) | Two organisms<br>f (%) | More than Two<br>organisms<br>f (%) |
| --- | --- | --- | --- |
| **Drinking water**<br>(n =15) | 3 (20.0) | 0 | 1 (6.7) |
| **Wastewater**<br>(n =15) | 12 (80.0) | 2 (13.3) | 1 (6.7) |

**Table 3:** Bacteria isolated from water samples.

| Name of organisms | Drinking water (n=15)<br>f (%) | Wastewater (n=15)<br>f (%) |
| --- | --- | --- |
| *Pseudomonas* spp. (n=9) | 3 (33.3) | 6 (66.7) |
| *Enterobacter* spp. (n=7) | 1 (14.3) | 6 (85.7) |
| *Escherichia coli* (n=5) | 0 | 5 (100.0) |
| *Citrobacter freundii* (n=1) | 0 | 1 (100.0) |
| *Citrobacter koseri* (n=1) | 0 | 1 (100.0) |
| *Acinetobacter* spp. (n=1) | 1 (100.0) | 0 |
| *Klebsiella oxytoca* (n=1) | 1 (100.0) | 0 |
| Total (n=25) | 6 (24.0) | 19 (76.0) |

**Table 4:** Biochemical test results for bacteria isolated from drinking water samples.

| Strain code | Biochemical tests |  |  |  |  |  |  |  |  | Isolated organism |
| --- | --- | --- | --- | --- | --- | --- | --- | --- | --- | --- |
|  | Catalase | Oxidase | TSI | Gas Production | H<sub>2</sub>S Production | Motility | Indole | Urease production | Citrate |  |
| DW06 | + | + | alk/alk | -- | -- | + | -- | -- | + | Pseudomonas spp |
| DW07 | + | + | alk/alk | -- | -- | + | -- | -- | + | Pseudomonas spp |
| DW09 | + | -- | aci/aci | +++ | -- | -- | -- | + | + | Klebsiella oxytoca |
| DW13 | + | -- | aci/aci | -- | -- | + | -- | -- | + | Enterobacter spp. |
|  | + | + | alk/alk | -- | -- | + | -- | -- | + | Pseudomonas spp |
|  | + | -- | alk/alk | -- | -- | -- | -- | -- | + | Acinetobacter spp |

**Table 5:** Biochemical test results for bacteria isolated from wastewater samples.

|  | Biochemical tests |  |  |  |  |  |  |  |  | Isolated organism |
| --- | --- | --- | --- | --- | --- | --- | --- | --- | --- | --- |
|  | Catalase | Oxidase | TSI | Gas Production | H<sub>2</sub>S Production | Motility | Indole | Urease production | Citrate |  |
| WW-01 | + | + | alk/alk | -- | -- | + | -- | -- | + | Pseudomonas spp |
| WW-02 | + | -- | aci/aci | +++ | -- | + | + | -- | -- | Esch coli |
|  | + | -- | aci/aci |  |  |  |  |  |  | Enterobacter spp. |
| WW-03 | + | -- | aci/aci | ++ | -- | + | -- | -- | + | Enterobacter spp. |
| WW-04 | + | + | alk/alk | -- | -- | + | -- | -- | + | Pseudomonas spp |
| WW-05 | + | + | alk/alk | -- | -- | + | -- | -- | + | Pseudomonas spp |
| WW-06 | + | + | alk/alk | -- | -- | + | -- | -- | + | Pseudomonas spp |
| WW-07 | + | -- | aci/aci | ++ | -- | + | + | -- | -- | Esch coli |
| WW-08 | + | -- | aci/aci | ++ | -- | + | + | -- | -- | Esch coli |
| WW-09 | + | + | alk/alk | -- | -- | + | -- | -- | + | Pseudomonas spp |
|  | + | -- | aci/aci | ++ | -- | + | + | -- | -- | Citrobacter freundii |
| WW-10 | + | -- | aci/aci | +++ | -- | + | -- | -- | + | Esch coli |
| WW-11 | + | -- | aci/aci | ++ | + | + | + | -- | + | Citrobacter koseri |
| WW-12 | + | + | alk/alk | -- | -- | + | -- | -- | + | Enterobacter spp. |
|  | + | -- | aci/aci | ++ | -- | + | + | -- | -- | Esch coli |
|  | + | + | alk/alk | -- | -- | + | -- | -- | + | Pseudomonas spp |
| WW-13 | + | -- | aci/aci | + | -- | + | -- | -- | + | Enterobacter spp. |
| WW-14 | + | -- | aci/aci | + | -- | + | -- | -- | + | Enterobacter spp. |
| WW-15 | + | -- | aci/aci | -- | -- | + | -- | -- | + | Enterobacter spp. |

### Antimicrobial susceptibility tests

The isolates were subjected to antimicrobial susceptibility tests by disc diffusion against 13 different antimicrobial agents in order to evaluate their resistance patterns. The phenotypic susceptibilities of the isolates against the different tested antimicrobial agents have been summarized in Table 6 (of drinking water) and Table 7 (of wastewater). The majority of the isolates exerted resistance to more than one group of antibiotics. The result of disc diffusion susceptibility testing revealed that most sensitive antimicrobials were meropenem, imipenem, gentamicin, ciprofloxacin, and amikacin.

**Table 6:** Antibiotic sensitivity test results for bacteria isolated from drinking water samples.

| Strain code | Name of organisms | Antibiotic sensitivity test |  |  |  |  |  |  |  |  |  |  |
| --- | --- | --- | --- | --- | --- | --- | --- | --- | --- | --- | --- | --- |
|  |  | AK | AMC | AMP | AT | CFM | CIP | CTR | CXM | GEN | IPM | MRP |
| DW6 | Pseudomonas spp | S | - | - | I | - | S | - | - | S | I | S |
| DW7 | Pseudomonas spp | I | - | - | R | - | S | - | - | S | I | S |
| DW9 | Klebsiella oxytoca | S | R | R | S | R | - | R | R | S | R | S |
| DW13 | Enterobacter spp. | S | R | - | - | R | S | R | R | S | S | S |
|  | Pseudomonas spp | R | - | - | S | - | S | - | - | R | I | S |
|  | Acinetobacter spp | R | R | - | S | - | S | R | - | R | R | S |

(AK-amikacin, AMC-amoxicillin with clavulanic acid, AMP-ampicillin, AT- aztreonam, CFM- cefuroxime, CIP- ciprofloxacin, CTR-ceftriaxone, CXM- cefixime, GEN- gentamicin, IPM- imipenem, MRP- Meropenem)

**Table 7:** Antibiotic sensitivity test results for bacteria isolated from wastewater samples.

| Strain code | Name of organisms | Antibiotic sensitivity test |  |  |  |  |  |  |  |  |  |  |  |  |
| --- | --- | --- | --- | --- | --- | --- | --- | --- | --- | --- | --- | --- | --- | --- |
|  |  | AK | AMC | AMP | AT | CIP | CFC | CFM | COT | CTR | CXM | GEN | IPM | MRP |
| WW-01 | Pseudomonas spp | S | -- | -- | I | I | R | -- | -- | R | -- | S | I | R |
| WW-02 | Esch coli | S | R | -- | S | R | -- | R | -- | R | R | S | I | S |
|  | Enterobacter spp. | S | R | -- | -- | R | -- | R | R | R | R | S | S | S |
| WW-03 | Enterobacter spp. | R | -- | -- | -- | R | -- | -- | R | R | R | R | R | R |
| WW-04 | Pseudomonas spp | S | -- | -- | I | R | R | -- | -- | R | -- | S | R | R |
| WW-05 | Pseudomonas spp | S | R | -- | R | R | R | -- | -- | R | -- | R | R | R |
| WW-06 | Pseudomonas spp | I | -- | -- | R | I | -- | -- | -- | R | -- | I | R | R |
| WW-07 | Esch coli | I | R | -- | -- | S | -- | -- | S | R | S | S | I | S |
| WW-08 | Esch coli | I | R | -- | -- | R | -- | R | T | R | R | R | R | S |
| WW-09 | Pseudomonas spp | R | I | -- | R | R | R | -- | -- | R | -- | R | R | R |
|  | Citrobacter freundii | S | R | -- | -- | S | -- | -- | R | R | R | R | -- | R |
| WW-10 | Esch coli | I | R | -- | S | I | -- | R | R | I | S | I | I | S |
| WW-11 | Citrobacter koseri | R | R | -- | -- | R | -- | -- | R | R | R | I | -- | R |
| WW-12 | Enterobacter spp. | S | R | -- | -- | R | -- | R | R | R | R | S | S | S |
|  | Esch coli | S | R | -- | S | R | -- | R | -- | R | R | S | I | S |
|  | Pseudomonas spp | S | R | -- | R | R | R | -- | -- | R | -- | R | R | R |
| WW-13 | Enterobacter spp. | R | R | -- | -- | R | -- | -- | R | R | R | R | I | S |
| WW-14 | Enterobacter spp. | S | R | -- | -- | S | -- | R | R | R |  | S | I | S |
| WW-15 | Enterobacter spp. | S | R | -- | -- | S | -- | -- | I | R | R | R | I | R |

(AK-amikacin, AMC-amoxicillin with clavulanic acid, AMP-ampicillin, AT- aztreonam, CIP- ciprofloxacin, CFC- cefepime with clavulanic acid, CFM- cefuroxime, COT- co-trimoxazole, CTR-ceftriaxone, CXM- cefixime, GEN- gentamicin, IPM- imipenem, MRP- Meropenem)

### Detection of resistance genes among drinking samples

The isolates were subjected to detect suspected ESBL and carbapenemase producing resistance genes by PCR in order to evaluate their resistance patterns. Table 8 shows distribution of two ESBL encoding genes (*bla*SHV and *bla*CTX-M-15) and five carbapenemase encoding genes (*bla*IMP, *bla*VIM, *bla*KPC, *bla*OXA1 and blaNDM1) among suspected resistant bacteria isolated from drinking water samples. Among four suspected ESBL producers, none were positive for ESBL encoding *bla*SHV and *bla*CTX-M-15 genes. Whereas among four suspected carbapenemase-producing bacteria, only one (25%) (*Pseudomonas* spp) had carbapenemase encoding genes *bla*KPC and others did not.

**Table 8:** Suspected Gene detection for resistant bacteria isolated from drinking water samples.

| Strain code | Name of organisms | Suspected Genes |  |  |  |  |  |  |
| --- | --- | --- | --- | --- | --- | --- | --- | --- |
|  | By Culture | SHV | CTX-15 | IMP | VIM | KPC | OXA1 | NDM1 |
| DW6 | Pseudomonas spp | X | X | X | X | X | X | X |
| DW7 | Pseudomonas spp | X | X | X | X | √ | X | X |
| DW9 | Klebsiella oxytoca | X | X | X | X | X | X | X |
| DW13 | Acinetobacter spp. | X | X | X | X | X | X | X |

### Detection of resistance genes among wastewater samples

The isolates were subjected to detect suspected ESBL and carbapenemase producing resistance genes by PCR in order to evaluate their resistance patterns. Table 9 shows distribution of two ESBL encoding genes (*bla*SHV and *bla*CTX-M-15) and five carbapenemase encoding genes (*bla*IMP, *bla*VIM, *bla*KPC, *bla*OXA1 and blaNDM1) among suspected resistant bacteria isolated from wastewater samples. Among 15 suspected ESBL producers, three (20%) and eight (53.3%) were positive for ESBL encoding *bla*SHV and *bla*CTX-M-15 genes respectively. On the other hand, among 15 suspected carbapenemase-producing bacteria, *bla*KPC genes were the most prevalent in 46.7% organisms whilst *bla*VIM, *bla*IMP and *bla*NDM1were the less common carbapenemase producers constituting 20%, 6.7% and 6.7% among suspected resistant bacteria respectively, and none was found *bla*OXA1 producer.

**Table 9:** Suspected Gene detection for bacteria isolated from wastewater samples.

| Strain Code | Name Of Organisms | Suspected Genes |  |  |  |  |  |  |
| --- | --- | --- | --- | --- | --- | --- | --- | --- |
|  | By Culture | SHV | CTX-15 | IMP | VIM | KPC | OXA1 | NDM1 |
| WW1 | Pseudomonas spp. | X | X | X | X | X | X | √ |
| WW 2 | Esch. Coli | X | √ | X | X | √ | X | X |
| WW 3 | Enterobacter spp, | X | √ | X | X | √ | X | X |
| WW 4 | Pseudomonas spp . | X | X | X | X | X | X | X |
| WW 5 | Pseudomonas spp . | X | X | X | √ | √ | X | X |
| WW 6 | Pseudomonas spp . | √ | X | X | X | √ | X | X |
| WW 7 | Esch Coli | X | √ | X | X | X | X | X |
| WW 8 | Esch Coli | √ | √ | X | X | X | X | X |
| WW 9 | Citrobacter Freundi | X | √ | X | √ | √ | X | X |
| WW 10 | Esch Coli | X | √ | X | X | X | X | X |
| WW 11 | Citrobacter Koseri | X | X | X | X | √ | X | X |
| WW 12 | Pseudomonas spp , | √ | √ | X | X | X | X | X |
| WW 13 | Enterobacter spp | X | √ | √ | √ | X | X | X |
| WW 14 | Enterobacter spp | X | X | X | X | X | X | X |
| WW 15 | Enterobacter spp | X | X | X | X | √ | X | X |

According to sanitary inspection risk score classification by WHO (2011), most of the drinking water samples score 4 (60%) out of 11 and categorized as intermediate risk by hazard score (Table 10).

**Table 10:** Specific diagnostic information for assessment of Risk.

| Sanitary Inspection Risk Score (Y=1/N=0) |  |  |  |  |  |  |  |  |  |  |  |  |  |
| --- | --- | --- | --- | --- | --- | --- | --- | --- | --- | --- | --- | --- | --- |
|  |  | 1 | 2 | 3 | 4 | 5 | 6 | 7 | 8 | 9 | 10 | 11 |  |
| SL NO. | ID NO. | Is there any point of leakage between source and reservoir ? | Is there any cover in reserve tank? | If there is a cover, is it damaged or inadequate to prevent contamination? | Distance between service point and reservoir ? | Are there any leak in the distribution network? | In the reservoir crack or eroded or unclean? | Is there any leakage in reserve tank? | Is there any sanitation within 10 meters of the pipeline (e.g. Latrines, sewers,<br>septic<br>tank or<br>burial<br>ground) | Is this reservoir cleaned within a month? | How often is the reservoir cleaned? | Is there any authority that inspects the Reserve Tank? | Total score (out of 11)<br>Y=1,<br>N=0 |
| 1 | DWR<br>01 | N | Y | N | 100 MT | N | Y | N | N | N | yearly | N | 4 |
| 2 | DWR<br>02 | N | Y | N | Y | N | N | N | Y | N | 6months<br>interval | N | 4 |
| 3 | DWR<br>03 | N | Y | N | N | N | N | N | N | N | 6m-yearly | N | 2 |
| 4 | DWR<br>04 | N | Y | NA | Y | NK | N | N | N | N | yearly | N | 3 |
| 5 | DWR<br>05 | N | Y | Y | 200 MT | N | N | N | N | N | 6months<br>interval | N | 4 |
| 6 | DWR<br>06 | N | Y | N | 200FT | N | N | N | N | N | yearly | NA | 3 |
| 7 | DWR<br>07 | N | Y | N | NA | N | NA | N | Y | N | yearly | N | 3 |
| 8 | DWR<br>08 | N | Y | N | NA | N | N | N | Y | N | 3months | Y | 4 |
| 9 | DWR<br>09 | N | Y | N | Y | N | N | N | Y | N | yearly | N | 4 |
| 10 | DWR<br>10 | N | Y | N | 1<br>METER | N | N | N | N | N | yearly | Y | 4 |
| 11 | DWT0<br>1 | N | Y | N | 100 MT | N | Y | N | N | N | yearly | N | 4 |
| 12 | DWT0<br>2 | N | Y | N | Y | NK | N | N | Y | N | 6months<br>interval | N | 4 |
| 13 | DWT0<br>3 | N | Y | Y | Y | N | N | N | N | N | N | N | 3 |
| 14 | DWS0<br>1 | N | N | Y | 200 MT | N | N | N | N | N | 6months<br>interval | N | 3 |
| 15 | DWS0<br>2 | N | Y | N | 200FT | N | N | N | N | N | yearly | Y | 4 |

**Table 11:** Sanitary inspection risk score classification.

| Hazard score | Risk interpretation | Findings n (%) |
| --- | --- | --- |
| 0-2 | Low risk | 2 (18.1) |
| 3-5 | Intermediate risk | 9 (90.9) |
| 6-8 | High risk | 0 |
| ≥ 9 | Very high risk | 0 |

## Discussion

This study presents the distribution of AMR organism with their resistance genes in drinking water, and wastewater in urban settings in Bangladesh. The high abundance of AMR organisms in wastewater within all settings, and drinking water in the urban area, raise important public health concerns.

In the present study, 63.3% of the water samples provided growths, while 26.7% of the drinking water and all of the wastewater samples yielded growths. Among them, 20%) of the drinking water samples yielded a single organism, whereas one (6.7%) sample yielded more than two organisms. The wastewater, on the other hand, yielded a single organism from 80.0% of samples, two organisms from 13.3% samples, and more than two organisms from one (6.7%) sample. Moges et al. (2014) reported that 85% of the samples were positive to one or more isolates which was higher than our study. They reported that among the total samples 113 bacterial isolates were recovered and 65 (57.5%) were from hospital environment and 48 (42.5%) were from non-hospital environment. ^49^ Similar findings found in a study conducted in Nepal showed the presence of heterotrophic bacteria in both tap and bottled water samples.^50^

In this study, 25 organisms were isolated from 30 samples of both drinking and wastewater and the predominant bacteria were found as *Pseudomonas* spp. (36%) followed by *Enterobacter* spp. (28%), *Escherichia coli* (20.0%), *Citrobacter freundii* (4.0%)*, Citrobacter koseri* (4.0%)*, Acinetobacter* spp.(4.0%) and *Klebsiella oxytoca* (4.0%) (Table 3). There are many studies that have shown the presence of different *Pseudomonas* spp. in drinking water.^51–53^ Similar findings of this study were reported in a study by Mulamattathil et al. (2014) in South Africa reported that *Pseudomonas* species were isolated in large numbers in raw and treated water. The study demonstrated the occurrence of total coliforms, faecal coliforms, heterotrophic bacteria, and *Pseudomonas* in water samples analysed which indicated the incidence of water contamination as some of these species are indicators of faecal contamination. ^54^

The members belonging to the family Enterobacteriaceae, including *E. coli* and *E. cloacae*, are known to be potential pathogens found in soil and water. Although *E. cloacae* is not a primary human pathogen, it has been reported to cause nosocomial infections.^55^

The presence of coliforms like *Escherichia coli* with other enteric bacteria showed faecal contamination of the water, which poses major health risks. Bhum*bla* et al (2020) from India reported that *Esch. coli* was the predominant bacteria along with *Klebsiella* spp. and Coagulase Negative *Staphylococcus* (CONS) respectively in the water which were not similar with the present study. ^56^

A study by Deji-Agboola et al. in Nigeria revealed the presence of coliforms in the tap water samples including *E. coli, Klebsiella spp.* and *Enterobacter spp.* which were similar to our study. ^57^ In contrast to our study findings, several researchers reported that most frequently isolated bacterium was *Klebsiella* spp., followed by *Pseudomonas* spp., *E. coli* and Citrobacter spp, and S. aureus (8.2%) which were not similar findings of this study.^49,58,59^

In a study by Narciso-da-Rocha et al. isolated different species of *Acinetobacter* from tap water w*hich* are considered opportunistic pathogens and very common in nature: soil, water, plants (including fruits and vegetables), animals and organic material in decomposition (sewage). They are also frequently found in water treatment systems and a well-known nosocomial pathogen that causes infections in hospitalized patients.^60^ In contrast to our findings Ranbir et al (2020) investigated the tap water quality of public toilets in Amritsar, Punjab, India and found that of the 25 isolates, 5 were of *Bacillus* spp., 4 were of *Pseudomonas* spp., as well as *E. coli* and *Enterobacter spp.*^58^

### Antimicrobial susceptibility tests

The isolates were subjected to antimicrobial susceptibility tests by disc diffusion against 13 different antimicrobial agents and the majority of the isolates exerted resistance to more than one group of antibiotics. The result of disc diffusion susceptibility testing revealed that most sensitive antimicrobials were meropenem, imipenem, gentamicin, ciprofloxacin, and amikacin. Different types of multiple antibiotic resistance patterns were observed amongst all the bacterial groups isolated from the various sites. In some cases, some of the isolates were resistance to up to ten different antibiotics.

This study also provides evidence of the prevalence of antibiotic-resistant bacteria in tap water samples of different localities, similar to previous studies. ^61–63^ Antibiotic resistance among bacterial species is spreading at an alarming rate and the lack of effective antibacterial drugs against them is a major concern.

A study by Mulamattathil et al. (2014) in South Africa revealed that None of the isolates were resistant to ciprofloxacin but a large proportion of the environmental isolates were resistant to erythromycin, followed by trimethoprim and amoxicillin.^54^ Moges et al. (2014) reported that multiple drug resistance was also common in gram negative isolates to commonly used antibiotics in the study area. All isolates of E. coli, Citrobacter spp. and Enterobacter spp. were 100% resistant to ampicillin. The overall resistance of gram-negative bacteria to ampicillin was 97%, followed by cephalotin 49%, cotrimoxazole 38%, tetracycline 37%, naldixic acid 36%, cefotaxime 33%; least resistant being ciprofloxacin 12% which was consistent to our study.^49^

Antibiotic resistance in bacteria is mainly because of the indiscriminate use of antibiotics by humans against various bacterial diseases and infections, animal husbandry, and agriculture. The vertical and horizontal transfer of antibiotic resistance genes from one bacterium to another of the same or a different genus through the process of conjugation, transformation and transduction may disrupt the microbial balance in favor of resistant bacteria. ^64^ In most developing countries, effluents are discharged from the sewer systems directly into rivers and lakes and a major source of contamination of the aquatic environment. These untreated effluents may contain AR bacteria, which disseminate and potentially transfer their resistance genes.^65^ In particular, wastewater from pharmaceutical plants could play a role in the selection of antibiotic-resistant bacteria in sewage.^67^

The presence of Extended Spectrum β-lactamases (ESBLs) and Metallo β-lactamases (MBLs) in the isolates from environmental samples has important implications for humans who depend on public water supply and sanitation facilities. Several studies have reported chromosomally encoded efflux mechanisms, abundant in gram negative bacteria and particularly in *Pseudomonas* spp., *Escherichia coli* and *Enterococcus* spp further contribute to raising the antibiotic resistance level and resistance genes in environmental samples. ^68,69^

In our study among suspected resistant bacteria isolated from drinking water samples, none were positive for ESBL encoding *bla*SHV and *bla*CTX-M-15 genes. Whereas among four suspected carbapenemase-producing bacteria, only one (25%) (*Pseudomonas* spp) had carbapenemase encoding genes *bla*KPC and others did not. On the other hand among 15 suspected carbapenemase-producing bacteria isolated from wastewater samples, *bla*KPC genes were the most prevalent in 46.7% organisms whilst *bla*VIM, *bla*IMP and *bla*NDM1were the less common carbapenemase producers constituting 20%, 6.7% and 6.7% among suspected resistant bacteria respectively, and none was found *bla*OXA1 producer. A study was conducted by Islam MA (2017) among wastewater samples from hospital-adjacent areas (HAR) and from community areas (COM), as well as public tap water samples, and found that 51 (71%) samples were positive for NDM-1-producing bacteria, as evidenced by phenotypic tests and the presence of the blaNDM-1 gene, compared to 5 of 41 (12.1%) samples from COM samples (P 0.001). Klebsiella pneumoniae (44%) was the predominant bacterial species among blaNDM-1-positive isolates, followed by Escherichia coli (29%), Acinetobacter spp. (15%), and Enterobacter spp. (9%).

These bacteria were also positive for one or more other antibiotic resistance genes, including blaCTX-M-1 (80%), blaCTX-M-15 (63%), blaTEM (76%), blaSHV (33%), blaCMY-2 (16%), blaOXA-48-like (2%), blaOXA-1 (53%), and blaOXA-47-like (60%) genes.12 In another study by Toleman (2012) found that, in Bangladesh, the blaNDM-1 gene were in 62% of environmental waters and in fermentative and non-fermentative gram-negative bacteria. Resistance genes and plasmid profiles for a subset of Escherichia coli strains was found in a study of extensively drug-resistant NDM-1– encoding bacteria in the environment, Dhaka, Bangladesh.13 The presence of NDM-1 β-lactamase-producing bacteria in environmental samples in New Delhi has important implications for people living in the city who are reliant on public water and sanitation facilities.14

The use of sanitary inspections combined with periodic water quality testing has been recommended in some cases as screening tools for fecal contamination. First introduced in 1991 and published in the World Health Organization (WHO) monitoring guidelines in 1993, sanitary inspections have become a common component of global water quality surveillance programs. They were developed to provide a rudimentary comparable method for quantifying risk factors that can contribute to microbiological contamination of water sources. Sanitary inspections include a simple visual assessment of typically around 11 risk factor questions, specific to the source type, which are answered with yes or no responses. Each risk factor question is weighted equally. The sum of all the questions answered “yes” is the sanitary inspection score (SIS). The higher the SIS value the higher the category of risk.^70^ According to sanitary inspection risk score classification by WHO, most of the drinking water samples score 4 (60%) out of 11 and categorized as intermediate risk by hazard score. In contrast of our study, Murei et al (2023) in India’s District Municipality identified as having the highest sanitary risk score in different water sources which were a cause for concern used by community members for domestic purposes.^71^ In the Southern Region of Ethiopia, from the sanitary condition survey, 57.6% of the water sources exhibited from intermediate-to very high-risk level which demonstrated the sanitary condition of water sources.^72^ These findings also help to fill highlighted gaps in research on the microbiological quality of wastewater sources.

## Conclusions

Our study findings provide an insight into the waterborne transmission of AMR by analyzing the distribution of ARB and ARGs in different aquatic environments in Bangladesh. Despite a greater awareness of environmental AMR, this has not yet been prioritized within environmental health policies or country-level national action plans, especially in LMICs. This study provides baseline information for further investigations into the prevalence of AMR bacteria in hospital and communal effluents and their spread into the aquatic ecosystem, representing potential threats to public health. Since these organisms may be vital to the safety and well-being of patients who are hospitalized as well as individuals who are susceptible to infection. Therefore, water treatment plant should be improved and sanitary measures should be practiced.

## Limitation

In this study, genotyping of AMR with public health importance were implied rather than multiple antimicrobial resistances (MAR) profiles.

## Recommendations

Improved sanitary measure with proper wastewater treatment plant should be practiced. To reduce the burden of AMR in the environment, future interventions are needed which include improved biosecurity practices, particularly waste disposal systems, along with improved water, sanitation and hygiene infrastructure in rural and in urban areas. Maintenance of proper sanitary conditions in toilets, cleaning of dustbins, proper air ventilation, and disinfection of floors and chlorination of water can decrease the chances of microbial contamination. In addition, touch-free flushing, taps and door-opening devices should be recommended to avoid pathogen transmission through contact. Materials used in the construction of water distribution systems should be carefully selected to prevent biofilm formation by microbes.

Mapping of AMR using quantitative microbiological data from connected environmental departments can provide valuable insights into an ecological setting and this could be an important step in addressing environmental dimensions of AMR through tailored mitigation measures.

## Data Availability

All data produced in the present work are contained in the manuscript

## Funding agency

Bangladesh Medical and Research Council (BMRC)

## Acknowledgement

This research study is attributed to the Water Supply and Sewerage Authority (WASA), Dhaka, Bangladesh. The authors are indebted to all the medical technologists of the laboratory of NIPSOM for their assistance in microbiology study.

